# Interactive effects of bacterial vaginal colonization and HIV on pregnancy outcomes: A Systematic Review and Meta-analysis

**DOI:** 10.1101/2024.09.18.24313939

**Authors:** Dismas Matovelo, Quinn Goddard, Paul Sabuni, Benson Kidenya, Jennifer Downs, Moke Magoma, Jeremiah Seni, Kathleen Helen Chaput

**Author notes:** Corresponding author: Dismas Matovelo.

## Abstract

**Background:** The independent impact of HIV and bacterial vaginal colonization on pregnancy outcomes has been documented and is compounded by the burden of rapidly escalating antimicrobial resistance. However, the interactive effect of HIV and lower genital tract bacterial colonization, on pregnancy outcomes has not been thoroughly studied and is examined in our study.

**Methods:** We performed a systematic review and meta-analysis to quantitatively assess the interaction between HIV and vaginal bacterial colonization and associations with birth weight and preterm birth. We searched Ovid MEDLINE, Ovid EMBASE, CINAHL, Scopus, Web of Science, Cochrane Library, African Journals Online, and PubMed databases to identify studies published up to December 31, 2023. We included observational reporting on vaginal colonization with bacterial pathogens stratified by HIV status that reported pregnancy outcomes. We followed the Preferred Reporting Items for Systematic Reviews and Meta-Analyses guidelines and used a modified Newcastle-Ottawa Scale to assess study quality. Meta-analysis was conducted using random-effects modeling in STATA Version 18. Pooled log-odds ratios were calculated. The study protocol was registered in PROSPERO(CRD42023485123).

**Results:** We selected 13 studies, involving 6,073 pregnant women, from 5,807 studies identified. The overall pooled prevalence of bacterial colonization was 26%(95%CI:17.3-37.4). There was no significant effect of HIV status and vaginal colonization on birth weight(OR=1.2, 95%CI:-2.57-2.20, *p*=0.88) and borderline increased odds of preterm birth (OR=2.64, 95%CI:-0.01-1.94, *p*=0.05). There was no significant association between HIV status and bacterial colonization(OR=1.08, 95%CI =-0.91-1.07), nor in antimicrobial resistance between pregnant women with HIV and those without.

**Conclusion:** Bacterial colonization is prevalent among pregnant women, but there is no clear evidence to suggest that HIV and bacterial colonization interact to affect birth weight or preterm birth. Research with large sample sizes, strict selection criteria, reliable and valid measurement, adequate control for confounding variables, and birthweight and gestational age at delivery assessment as continuous outcomes are still needed to provide robust evidence.

## INTRODUCTION

Bacterial genital infections are usually polymicrobial, affecting up to 50% of women during pregnancy, intrapartum, and postpartum, and vary across trimesters due to factors such as hormonal changes, host genetics, behavioural, nutritional, and environment[1–3]. However, despite hormonal changes during pregnancy, vaginal bacterial composition during gestation typically remains relatively stable among pregnant women not living with HIV[4]. In the setting of HIV infection, which is characterized by an increase in pro-inflammatory responses and the release of cytokines, the integrity of the vaginal mucosal epithelium, the primary defence mechanism is disrupted. Due to this loss in integrity of the vaginal mucosal epithelium, the vaginal mucosa in HIV-infected pregnant women is characterized by a diverse microbiota predominantly comprised of anaerobic species[3].

The occurrence of lower genital tract infections in pregnancy has significantly been reduced in high-income countries (HICs) in recent decades, due to optimization of surveillance and treatment[5]. However, such routine surveillance for common pathogens colonizing the vagina during pregnancy lack in most Low-Middle Income Countries (LMICs), along with the rapidly escalating burden of antimicrobial resistance (AMR) and variations in worldwide resistance patterns. As a result, empirical treatment has become difficult in LMICs[6, 7].

Approximately 70% of people living with HIV are in sub-Saharan Africa, and the global prevalence of HIV among pregnant women is 7.2%[8, 9]. The dual impact of HIV infection and bacterial genital colonization may affect placental development and function, causing low birthweight (LBW), small for gestational age (SGA), intrauterine growth restriction (IUGR), or preterm birth (PTB). Over the past decade it has become clear that although a healthy placenta has no microbiome, it is still susceptible to pathogenic bacterial colonization, commonly group B Streptococcus[10]. Placental bacterial colonization can affect placental development and function, thereby impacting fetal growth and development. Disrupted placental function can lead to SGA, IUGR, preterm labour and birth, rupture of membranes (ROM), chorioamnionitis, neonatal sepsis, and LBW [10, 11]. These factors contribute to a series of short and long-term adverse effects such as neonatal sepsis and delayed child developmental milestones[10, 11].

Both HIV and bacterial genital colonization may have an impact on pregnancy outcomes, especially birthweight, which is an important health indicator that has long-lasting impacts on infant and childhood development[11]. Globally, LBW and PTB are common, affecting an estimated 19.8 and 13.4 million births in 2020, respectively, with increased mortality risks particularly in the neonatal period, and long-term adverse health outcomes throughout life[12, 13]. However, there is a paucity of evidence on the association between HIV infection, lower genital tract pathogenic bacterial colonization and birth weight and conflicting evidence for associations and potential interactions between bacterial colonization and HIV status with birthweight.

Several individual studies attempting to estimate the disease burden of lower genital tract bacterial colonization have reported that regardless of HIV status, *Escherichia coli, Klebsiella. pneumoniae* and *GBS* are the most common organisms, with a wide prevalence range of1.5%-73.5%[14, 15]. While some studies report a higher prevalence rate among pregnant women infected with HIV[16], compared with those without, others report no differences[17–19]. To bridge this knowledge gap, we performed a systematic review and meta-analysis to quantitatively assess the prevalence of lower genital tract bacterial infections and their association with pregnancy outcomes in HIV-infected and HIV-uninfected individuals. The results of this study provide insights into designing large prospective studies to decipher the associations between lower genital tract bacteria colonization, HIV infection, and maternal and perinatal outcomes. It will also help healthcare policymakers and various stakeholders ideally, seeking to standardize protocols for medical care and screening in this vulnerable population, with the ultimate goal of reducing associated adverse maternal and perinatal outcomes.

## METHODS

### Objective

The current review aimed to examine the interactive effects of lower genital tract bacterial colonization in pregnancy and HIV infection on the primary outcomes of birthweight and preterm birth. The secondary outcomes were intrauterine growth restriction (IUGR) and small for gestation age (SGA), as well as assessed the prevalence of antimicrobial resistance (AMR) among HIV-infected vs -uninfected pregnant women as a tertiary outcome. This review was conducted following PRISMA guidelines (S2 Appendix) [20] and was registered with the International Prospective Register of Systematic Reviews (PROSPERO)[21], registration number CRD42023485123, December 2023.

### Search Strategy

We identified studies using electronic research databases and hand-searching methods. The comprehensive search strategy was developed in consultation with a research librarian and reviewed by two perinatal researchers (S1 Appendix). Our controlled vocabulary search included the following exploded keywords: *Vaginitis, “Genital Diseases, Female”, “Reproductive Tract Infections”, vagina, bacteria, Escherichia coli, “Klebsiella pneumoniae”, “streptococcal infections”, HIV, HIV Seropositivity, HIV Infections, HIV Seronegativity, Pregnancy, Pregnant Women, Pregnancy Trimester, Third, and pregnancy outcome*.

We applied the search strategy in MEDLINE (Ovid), EMBASE (Ovid), CINAHL, Scopus, Web of Science and the Cochrane Library to identify studies published regardless of year. We also searched African Journals Online and PubMed databases for additional publications and used Web of Science, Google and Google Scholar to search for grey literature. We modified indexing terms such as Medical Subject Headings (MESH) or field tags according to each database and linked searches using “AND” and “OR” Boolean operator terms as required. We hand-searched by scanning references, “cited-by”, and “similar articles” lists for eligible studies. No filters were applied.

### Eligibility criteria

We initially included all quantitative observational studies that reported the prevalence of bacterial vaginal colonization published in English in peer-reviewed journals. Observational studies were included in the meta-analysis, that employed cohort, case-control, or cross-sectional designs. We included grey literature and published abstracts for a narrative summary in the review.

Eligible studies involved pregnant women of any age who were tested for vaginal bacterial colonization at any point during their gestation. Studies must have had an exposure arm with pregnant women with HIV infection and a control arm with those without HIV. Studies must have measured birth weight and/or preterm birth as a pregnancy outcome, to document the differences in associations between vaginal bacterial colonization and birth weight among pregnant women with and without HIV. The outcomes involved aggregate counts for preterm birth (< 37 weeks completed gestation) and low birth weight (< 2500g).

### Study selection

We used Covidence software to collate our initial search results and remove duplicates before screening. Two independent authors (DM and QG) screened titles and abstracts of each article and assigned a ‘yes’, ‘no’, or ‘maybe’ to their eligibility for inclusion. Studies marked as “yes” or “maybe” underwent a full-text review, during which reasons for exclusion were documented. Any conflicts in decision-making were resolved by a third author (KHC). The Kappa index of the inter-observer agreement for title/abstract and full-text selection was high (0.96 and 0.73, respectively) as calculated by Covidence.

### Data Extraction and Critical Appraisal

Authors (DM and QG) conducted data extraction using Covidence, which was then exported into Microsoft Excel 2023. We extracted the following variables from the articles: a) first author’s name, b) publication year, c) region/country, d) study type/design, e) type of sample collected, f) sample size, g) sampling method, h) the prevalence of vagina colonization, i) the microorganisms involved, j) pregnancy outcomes, and k) any other results relevant to primary and secondary outcomes (if applicable). Two authors (DM and QG) conducted the data extraction independently. Any discrepancies that emerged were resolved by a third individual (KHC). If desired data were available for extraction, we contacted authors a minimum of three times between January and June 2024 to obtain missing data points. Studies were excluded from the meta-analysis if the authors were unreachable or unable to share the required data.

### Quality assessment

We used the modified Newcastle-Ottawa Scale (NOS) for quality assessment since all eligible studies were observational designs[22]. We adapted the tool by adding topic-specific criteria to make it more relevant to an epidemiological context. Specifically, we added criteria for controlling additional confounders (such as co-morbidities), defining key variables (such as vaginal colonization), and reporting sample, exposure, and control group details.

Quality assessment was performed by the first (DM) and second (QG) authors, and verified by the senior author (KHC). The NOS scale consists of eight sections that evaluate an article based on selection criteria, comparability, exposure assessment, and outcomes. Each included study received a score corresponding to the number of quality criteria satisfied, with scores of 0-5 considered low quality, 6-7 considered moderate quality, and 8-10 considered high quality. We included all articles in the systematic review but only studies with a minimum score of 6 and above were included in the meta-analysis.

### Data analysis

A narrative analysis was performed to summarize the characteristics of the included studies. We quantified the pooled interactive effect of bacterial vaginal colonization and HIV on pregnancy outcomes, as well as the association between HIV and bacterial colonization, using random-effects maximum likelihood forest plots to calculate log-odds ratios (OR) and their corresponding 95% confidence intervals. We further quantified the interaction using Cochrane’s Q value and Higgins I-squared (I^2^)[23], with a continuity correction of 0.5 for insufficient cell sizes[23]. We categorized I^2^ values of 0–40% as low heterogeneity; 50–70% as moderate heterogeneity, and >70% as high heterogeneity[22]. The heterogeneity of studies reporting primary outcomes was further assessed using a Galbraith Plot with 95% confidence intervals. Publication bias was represented graphically using a contour-enhanced funnel plot. We conducted all analyses using Stata version 17.0.

## Results

We initially identified 7,005 identified studies (Medline = 981, Embase = 1,137, Web of Science = 276, CINAHL = 4,075, Scopus = 459, Cochrane = 61, and grey literature (ProQuest) = 14; 1,198 duplicates were excluded). The first and second authors screened the 5,805 remaining abstracts, deeming 5,719 irrelevant. This left 86 studies for assessment. Two studies were identified through hand-searching reference lists and were included in a full-text review (S1 Figure).

**S1 Figure:**
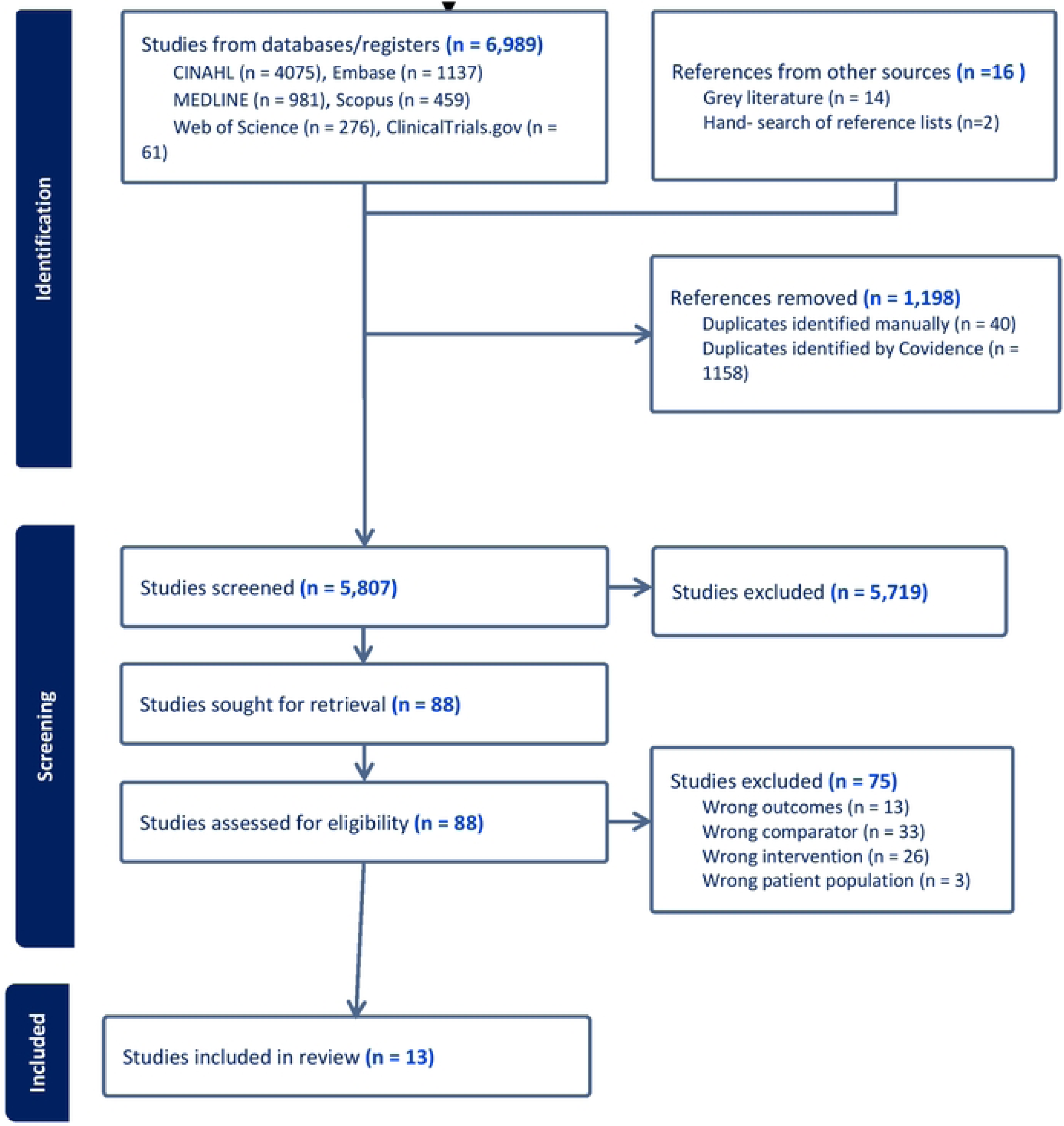
PRISMA flowchart diagram of the study selection.

The two authors had high inter-rater reliability (proportionate agreement = 0.97) and resolved 177 conflicting assessments through consensus. After a full-text review of 88 studies, 13 studies that met the inclusion criteria were included in the systematic review (Table in S1 Table). All identified studies were rated as 7 or higher by the first and second authors on the modified NOS. Therefore, no studies were excluded from the meta-analysis based on quality assessment (S2 Table).

**S1 Table:**
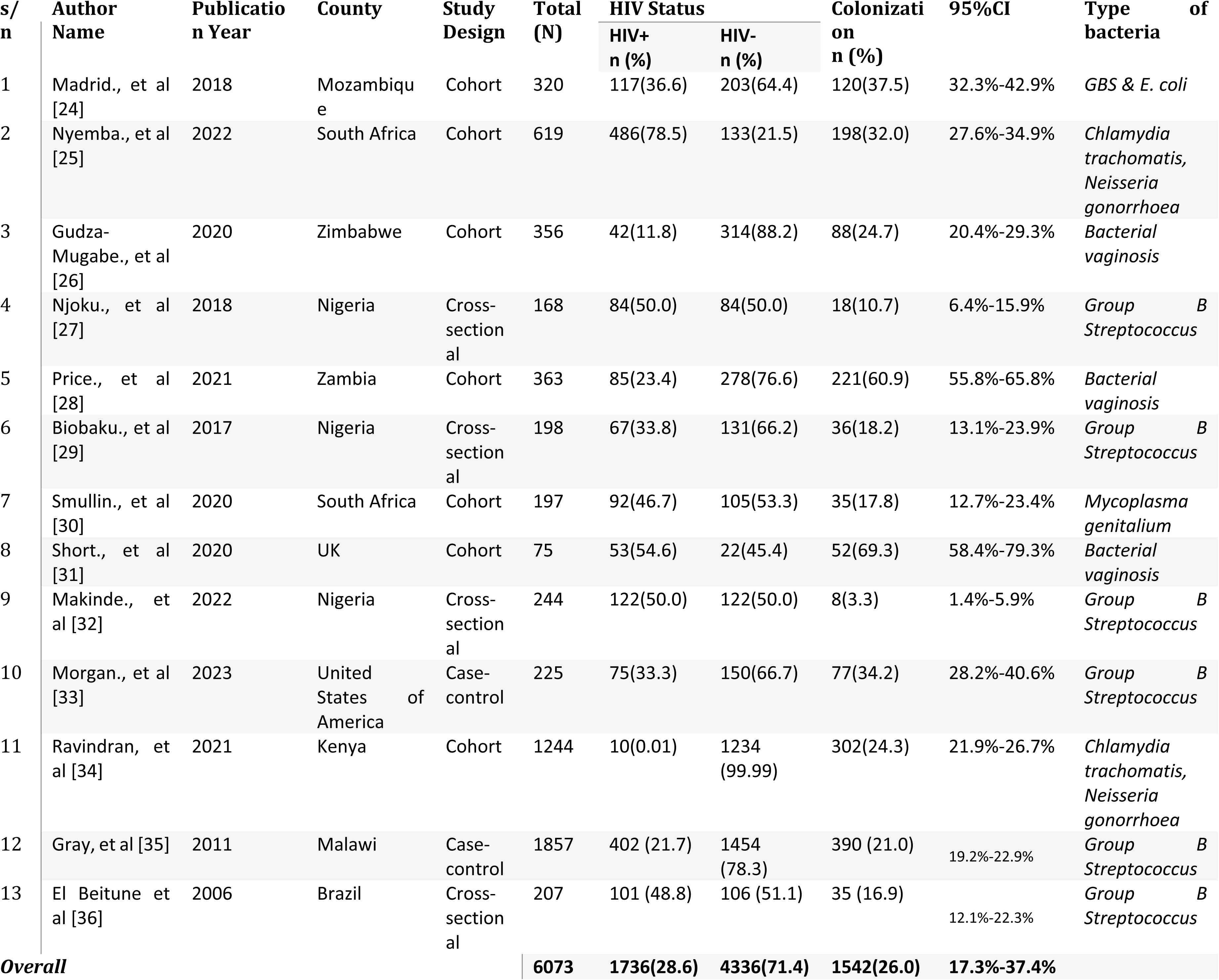
Characteristics of the included studies.

Since the majority of the included studies did not report all data points required for meta-analysis, we contacted the authors from all studies to request additional data. All included studies were conducted between 2006 and 2023 and published online between 2017 and 2023. Of the studies selected, four were cross-sectional, seven were cohort studies, and two were case-control studies. All except three were conducted in Africa. We found no other published systematic reviews on this topic.

The overall sample size of all included studies was 6,073 participants, with individual study sample sizes ranging from 75 to 1,857. The prevalence of vaginal bacterial colonization ranged from 3.3% to 69.3% (Table in S1 Table). Testing and diagnostic criteria for bacterial colonization varied (S2 Table).

**S2 Table:** Distribution of vaginal bacterial colonization among pregnant women.

| *Author name* | *Total (N)* | *Lab Method* | *Bacteria* | *HIV+ n (%)* | *95% CI* | *HIV- n (%)* | *95% CI* | *NOS* |
| --- | --- | --- | --- | --- | --- | --- | --- | --- |
| *Madrid (2018)* | 120 | Culture | *E.coli* | 44(36.7) | 28.2%-45.5% | 76(63.3) | 54.5%-71.8% | 8 |
| *Nyemba (2022)* | 198 | Culture | *Chlamydia trachomatis, Neisseria gonorrhoea* | 163(82.3) | 76.7%-87.3% | 35(17.7) | 12.7%-23.3% | 8 |
| *Gudza-Mugabe (2020)* | 88 | PCR & DNA Sequencing | *Bacterial vaginosis* | 18(20.5) | 12.6%-29.6% | 70(79.5) | 70.4%-87.4% | 7 |
| *Njoku (2018)* | 18 | Culture | *Group B Streptococcus* | 13(72.2) | 48.9%-90.9% | 5(27.8) | 9.1%-51.1% | 8 |
| *Price (2021)* | 221 | Metagenomic Sequencing | *Bacterial vaginosis* | 85(38.7) | 32.1%-45.0% | 136(61.5) | 55.0%-67.9% | 8 |
| *Biobaku (2017)* | 36 | Culture | *Group B Streptococcus* | 13(36.1) | 21.1%-52.6% | 23(63.9) | 47.4%-78.9% | 10 |
| *Smullin (2020)* | 35 | Culture | *mycoplasma genitalium* | 22(62.9) | 46.1%-78.2% | 13(37.1) | 21.8%-53.9% | 8 |
| *Short (2020)* | 52 | DNA Sequencing | *Bacterial vaginosis* | 42(80.8) | 68.8%-90.5% | 10(19.2) | 9.5%-31.2% | 7 |
| *Makinde (2022)* | 8 | Culture | *Group B Streptococcus* | 4(50.0) | 15.3%-84.7% | 4(50.0) | 15.3%-84.7% | 8 |
| *Morgan (2023)* | 77 | Culture | *Group B Streptococcus* | 31(40.3) | 29.5%-51.5% | 46(59.7) | 48.5%-70.5% | 7 |
| *Ravindran (2021)* | 302 | Culture | *Chlamydia trachomatis, Neisseria gonorrhoea* | 4(0.01) | 0.0%-3.0% | 296(99.99) | 97.0%-99.7% | 8 |
| *Gray (2011)* | 390 | Culture | *Group B Streptococcus* | 77(19.4) | 15.9%-23.8% | 313(21.7) | 76.2%-84.1% | 7 |
| *El Beitune (2006)* | 207 | culture | *Group B streptococcus* | 20(19.8%) | 40.3%-73.2% | 15(14.1%) | 26.8%-59.7% | 7 |

### Narrative analysis

No clear pattern of association between HIV, bacterial colonization, and low birth weight was observed in the included studies. For instance, Makinde *et al*[32] found no association between vaginal colonization and HIV status regarding low birth weight. Potentially, the very low prevalence of GBS within their sample (8/244), leading to low power in detecting an effect may explain the overall findings of this study. Similarly, Gudza-Mugabe *et al*[26] found no interaction between vaginal microbiota and HIV positivity in determining birth outcomes. In contrast, Nyemba *et al*[25] found that the presence of *C. trachomatis* and *N. gonorrhoea* at the first antenatal care (ANC) visit in individuals with HIV leads to an increased risk of an adverse birth outcome. It is important to note that the the bacterial species examined varied across studies (i.e., *GBS*, *C. trachomatis, N. gonorrhoeae*).

Results for preterm birth were more consistent across the included studies. Nyemba *et al*[25], Smullin *et al*[30], and Short *et al*[31] all found an elevated risk of preterm birth in analyzing the interaction between vaginal colonization and HIV status. However, Gudza-Mugabe *et al*[26] and Price *et al*[28] found that individuals with HIV infection were more likely to give birth prematurely, regardless of vaginal microbiota, suggesting that the relationship between HIV status and preterm birth was not modified by bacterial colonization. Ravindran *et al*[34] found that incident HIV infection, as well as *C. trachomatis* and *N. gonorrhoeae* infection, all independently increased the risk of PTB. El Beitune *et al*[36] found no significant difference in the prevalence of GBS carriage between HIV-positive and HIV-negative participants as well as no significant difference in gestational age and birthweight of infants born to HIV-positive mothers compared with those born to HIV-negative mothers. The study did not stratify birth outcomes by *GBS* status. Since most of these papers did not stratify by bacterial colonization status and HIV status, the data included in our meta-analysis were obtained directly from the authors upon request.

Benedetto *et al*[37] tested for BV as well as *C. trachomatis*, but did not stratify birth outcome by HIV status with regards to colonization. The study found that cervicovaginal infections increased the chance of preterm delivery. Foessleitner *et al*[38] investigated vaginal dysbiosis and BV in HIV-positive and HIV-negative women. While preterm birth was equally likely regardless of HIV status, it was found that HIV-positive women were more likely to develop dysbiosis or BV. Govender *et al*[39] analyzed the prevalence of genital mycoplasmas, ureaplasmas, and *Chlamydia trachomatis* to HIV status. There was no association between HIV status and bacterial colonization, as well as preterm delivery and colonization.

Several additional studies examined our variables of interest, but a lack of access to the original dataset (e.g. destruction or loss of data) meant we were unfortunately not able to include them in our meta-analysis. Zenebe *et al*[40] documented STI status during pregnancy but did not analyze the interaction between STI and HIV status on pregnancy outcomes. Looking at HIV individually, they found that the mean birth weight was higher in babies born to HIV-uninfected women, and women with HIV were more likely to give birth to a LBW infant than those without. Joachim *et al*[41] examined GBS colonization as well as HIV status, finding no significant association between infant birthweight and GBS carriage, but did not examine differences by HIV status. Seale *et al*[42] found that women with GBS had a higher chance of stillbirth, but equal likelihood of very preterm birth and very low birthweight than those without. It was found that GBS carriage was reduced overall in women with HIV, but that women with HIV were more likely to have more virulent CC17 GBS. Temmerman *et al*[43] tested for BV, *C. trachomatis, N. gonorrhoeae*, as well as HIV. It was found that mothers with HIV were more likely to give birth to small for gestational age infants. Madrid *et al*[24] tested for *Group B Streptococcus* and found that mothers with HIV were at no higher risk of *GBS* colonization than those without. Morgan *et al*[33] found that HIV-positive individuals had a higher chance of preterm birth, but that the likelihood of GBS colonization did not differ between individuals with and without HIV.

Only one study, Gudza-Mugabe *et al*[26], analyzed small for gestational age (SGA), and found no difference in SGA between mothers with and without HIV. None of the included studies analyzed intrauterine growth restriction.

### Meta-Analysis

#### Primary Outcomes

Of the studies with adequate data to be included in the meta-analysis, five examined low birth weight as an outcome, with an overall sample of 3,520 individuals, 784 (22.2%) of whom had bacterial colonization and 1,284 (36.4%) were pregnant women with HIV infection. Six studies examined preterm birth, with a total sample of 5,164 individuals, 1,356 (26.3%) of which had bacterial colonization and 1,372 (26.6%) were pregnant women with HIV.

##### Low Birthweight

We observed no significant interactive effect between HIV status and vaginal bacterial colonization on low birth weight (log OR= 0.19, 95% CI= −2.57,2.20, equivalent OR=1.21, *p* = 0.88) and a high degree of heterogeneity between the studies (*I^2^*= 88.73%; *Q*= 19.84, *p*= 0.00), indicating low reliability for the pooled estimate (S2 Figure).

**S2 Figure:**
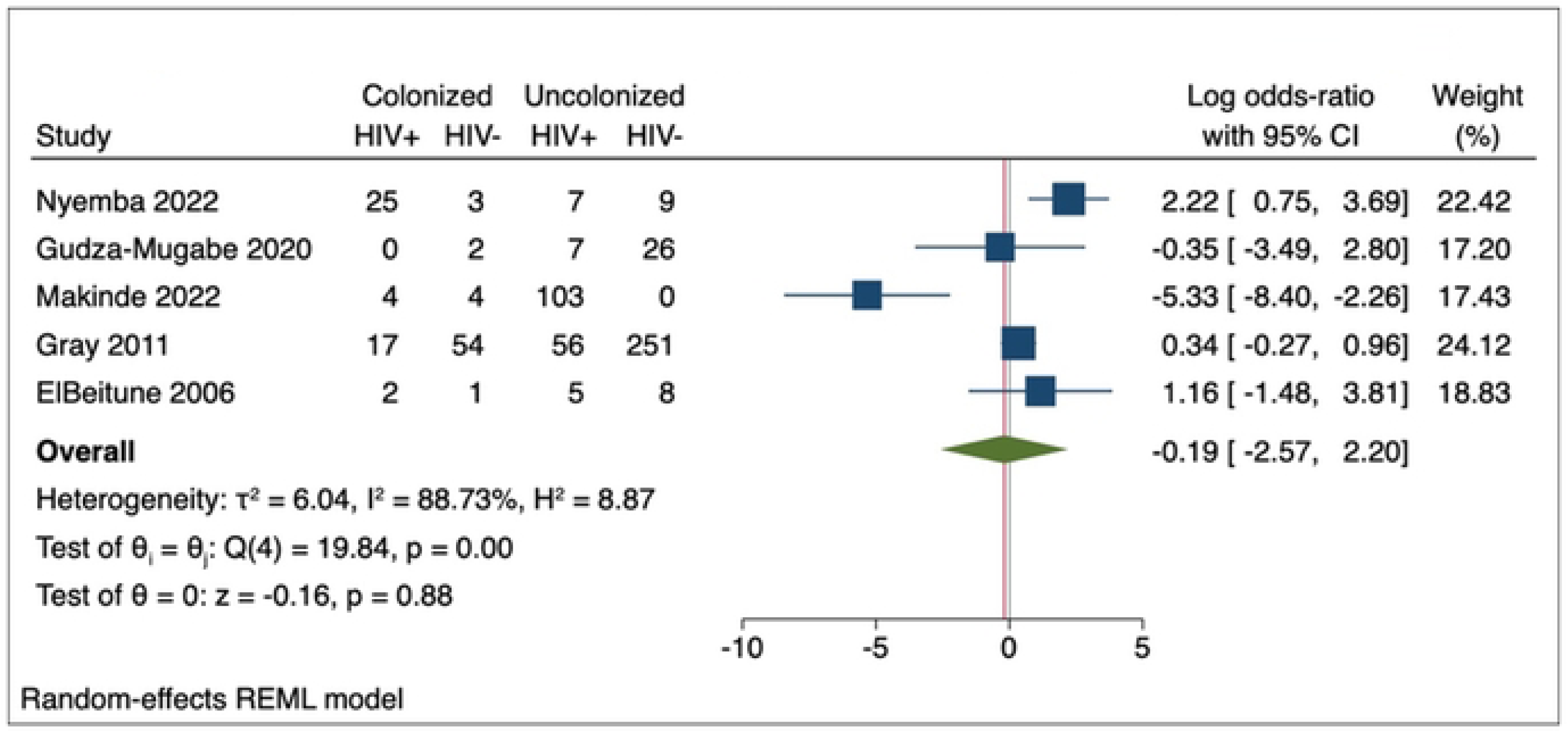
Interactive effect of HIV and bacterial colonization on birthweight.

##### Preterm Birth

Overall, we found a borderline-significant increased log-odds of preterm birth among individuals with HIV and vaginal bacterial colonization, compared to those without either (log OR= 0.97, 95% CI=-0.01, 1.94, *p*=0.05; equivalent OR: 2.64, *p* = 0.05). Moderate heterogeneity was observed between the included studies (*I^2^* = 53.76%; *Q* = 16.78, *p* = 0.02), indicating moderate reliability of the estimate. One study [34] reported a log-OR less than 0 (indicating the protective effect of HIV and Colonization), while the majority of studies (75%) trended toward increased odds of preterm birth with HIV and bacterial colonization compared to no HIV or colonization (S3 Figure).

**S3 Figure:**
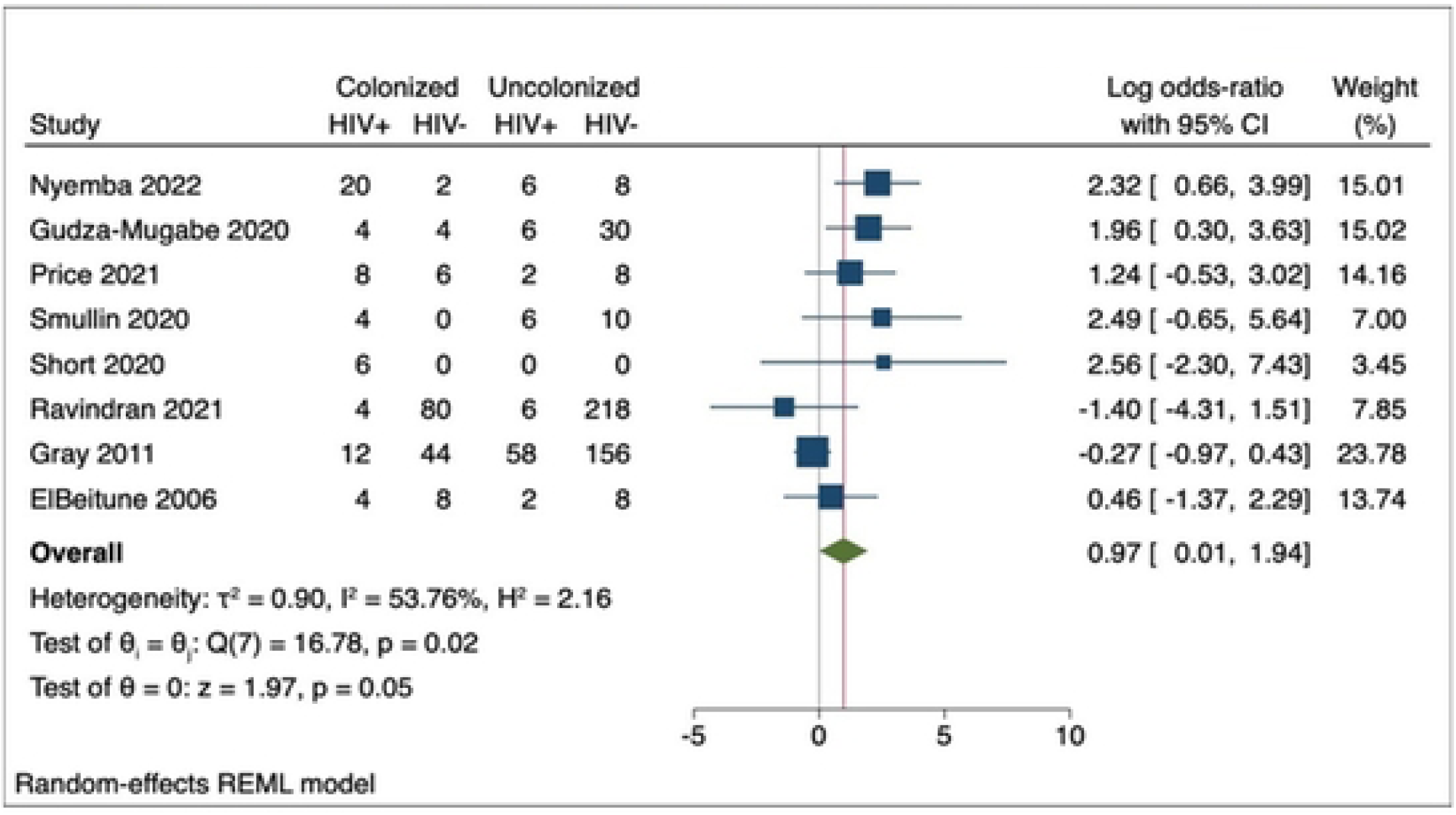
Interactive effect of Hiv and bacterial colonization on preterm birth.

The methods of calculating gestational age varied, potentially impacting the estimate of preterm birth. Three studies [25, 26, 35] used an estimate of last menstrual period (LMP), one study [28] used ultrasound dating, and one study [34] employed a combination of ultrasound, LMP and fundal height. The two remaining studies [30, 31] did not specify the method of GA calculation.

### Heterogeneity

We lacked enough studies in the pooled analysis of low birthweight to effectively evaluate heterogeneity or publication bias graphically for this outcome. The Galbraith plot of preterm birth studies confirms overall low-moderate heterogeneity, with all 8 studies lying within the 95% CI region (S4 Figure). The regression line significantly deviates from the null-effect line, with 75% of the studies positioned above the no-effect line.

**S4 Figure:**
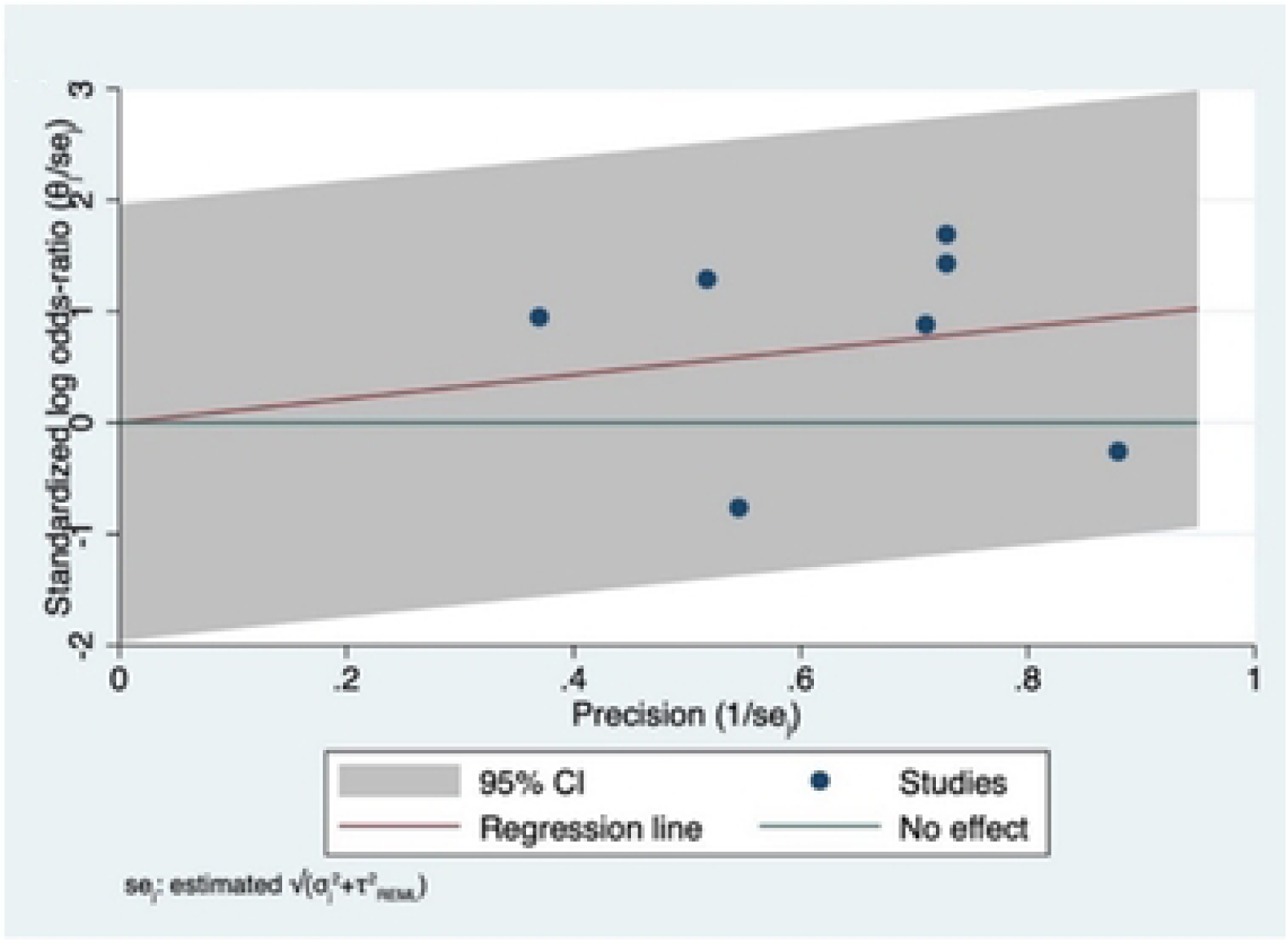
Galbraith plot of precision in studies examining low birthweight.

### Publication Bias

The included studies produce a right-skewed, asymmetrical funnel (S5 Figure), suggesting the presence of publication bias. This bias likely stems from the underreporting of null or protective findings, potentially leading to an overestimation of the effect in the included studies. Therefore, although 7 out of 8 studies lie within the 90% confidence interval, these findings should be interpreted cautiously due to the relatively low number of studies included in this assessment (<10).

**S5 Figure:**
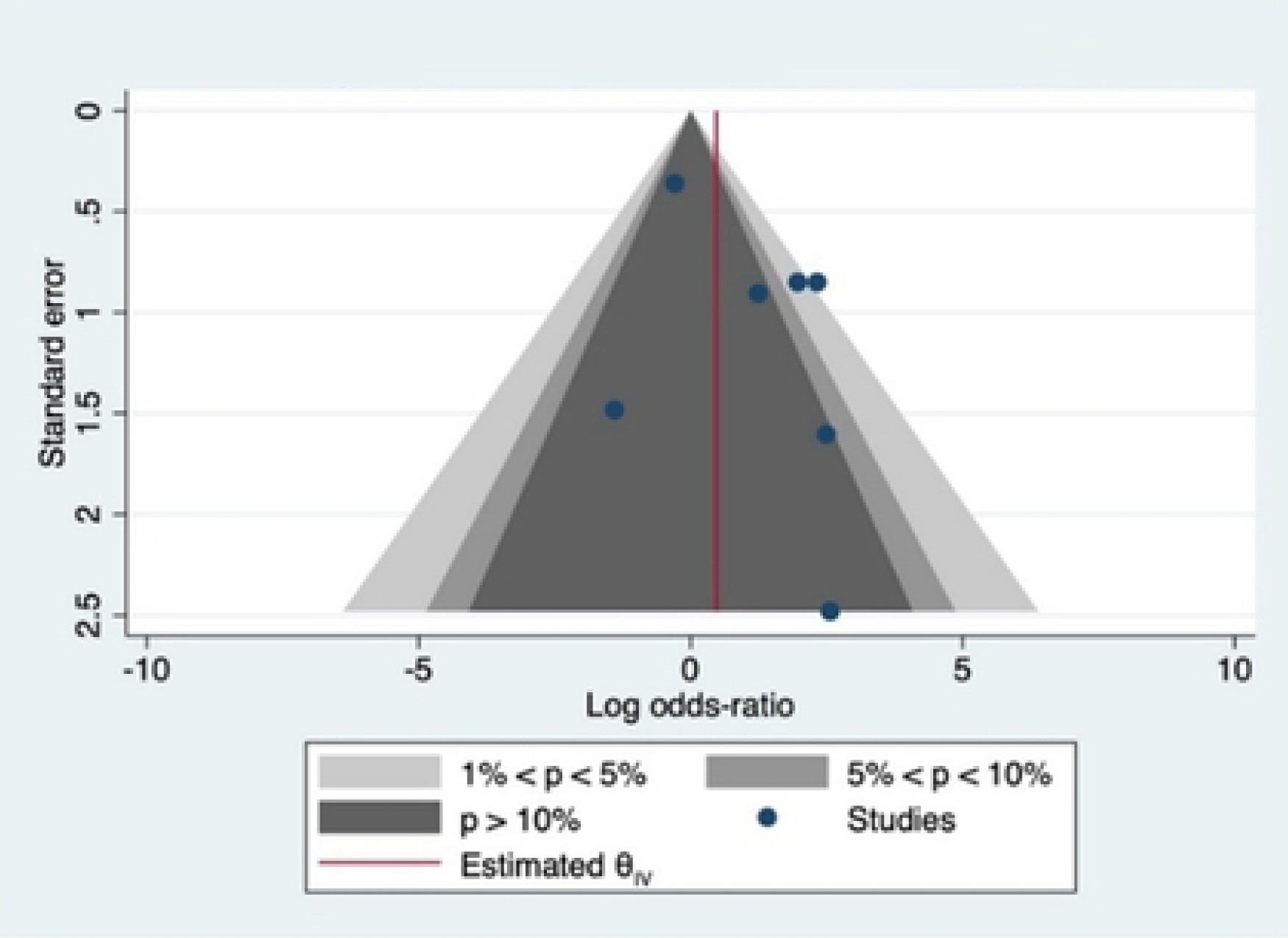
Publication bias contour-enhanced funnel plot.

#### Secondary Outcomes

The birth outcomes examined in the included studies varied considerably, and we had insufficient data to generate meaningful pooled estimates for our secondary outcomes of intrauterine growth restriction (IUGR), and small for gestational age (SGA). This was defined as a birth weight of less than the 10^th^ percentile for gestational age.

#### Tertiary Outcome

In assessing antimicrobial resistance (AMR), we found that six different types of bacterial isolates were examined in the included studies, with the highest pooled prevalence being bacterial vaginosis (33.9%), examined in three studies. Six of the included studies tested for Group B Streptococcus (GBS) which had a pooled prevalence of 20.2%. Other bacterial species were less commonly examined (S3 Table). All studies testing AMR evaluated *GBS*, facilitating meaningful pooled estimates of the association between HIV and *GBS* in terms of AMR.

**S3 Table:** Vaginal colonization bacterial isolate types.

| *Bacterial isolates* | *No. of studies* | *Total N* | *Cases* | *Prevalence, 95% CI* |
| --- | --- | --- | --- | --- |
| *Group B Streptococcus* [27, 29, 32, 33, 35, 36] | 6 | 3129 | 632 | 20.2%, 18.8%-21.6% |
| *Mycoplasma genitalium* [30] | 1 | 197 | 35 | 17.8%, 12.7%-23.4% |
| *Bacterial vaginosis* [26, 28, 31] | 3 | 794 | 269 | 33.9%, 30.6%-37.2% |
| *Neisseria Gonorrhoea* [25] | 1 | 619 | 35 | 5.7%, 4.0%-7.6% |
| *Chlamydia trachomatis* [25] | 1 | 619 | 158 | 25.5%, 22.2%-29.0% |
| *Escherichia coli* [24] | 1 | 320 | 52 | 16.2%, 12.4%-20.5% |
| *Any STI in pregnancy* [25, 30] | 2 | 816 | 311 | 38.1%, 34.8%-41.5% |
| *Chlamydia trachomatis & Neisseria gonorrhoea* [34] | 1 | 1244 | 302 | 24.3%, 21.9%-26.7% |

There was no significant association between HIV status and bacterial colonization across included studies (log-OR=0.08 [-0.91, 1.07]; equivalent OR=1.08) (S6 Figure). Likewise, we found no statistically significant difference in log odds of AMR between HIV-positive and HIV-negative individuals for each antibiotic examined for resistance (Ampicillin: log-OR=0.47[-0.94, 1.87]; equivalent OR=1.60, Ceftriaxone: log-OR=0.28[-1.77, 2.33]; equivalent OR=1.32, Clindamycin: log-OR=0.48[-1.28, 2.24], equivalent OR=1.61, Erythromycin: logOR=1.14[-0.83, 3.11], equivalent OR=3.12, Penicillin: log-OR=0.42 [-0.94, 1.77], equivalent OR=1.42). Heterogeneity was low to moderate for most antibiotics, except for erythromycin (I^2^=75.79). (S7 Figure: A-E) We were unable to pool proportions of antimicrobial resistance across antibiotic types, due to aggregate reporting of AMR, and lack of individual participant data.

**S6 Figure:**
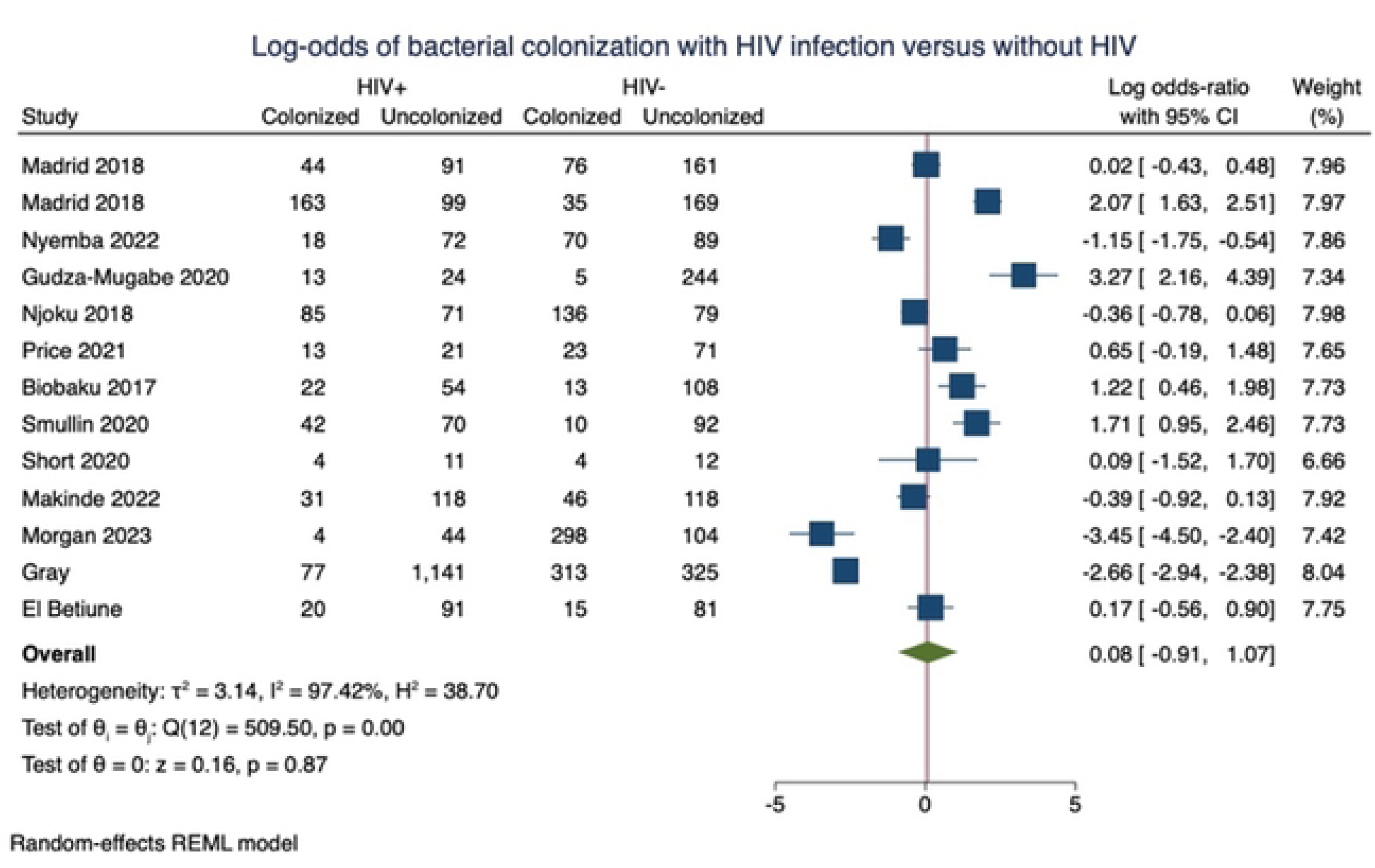
Association between HIV status and Bacterial Colonization.

**S7 Figure:**
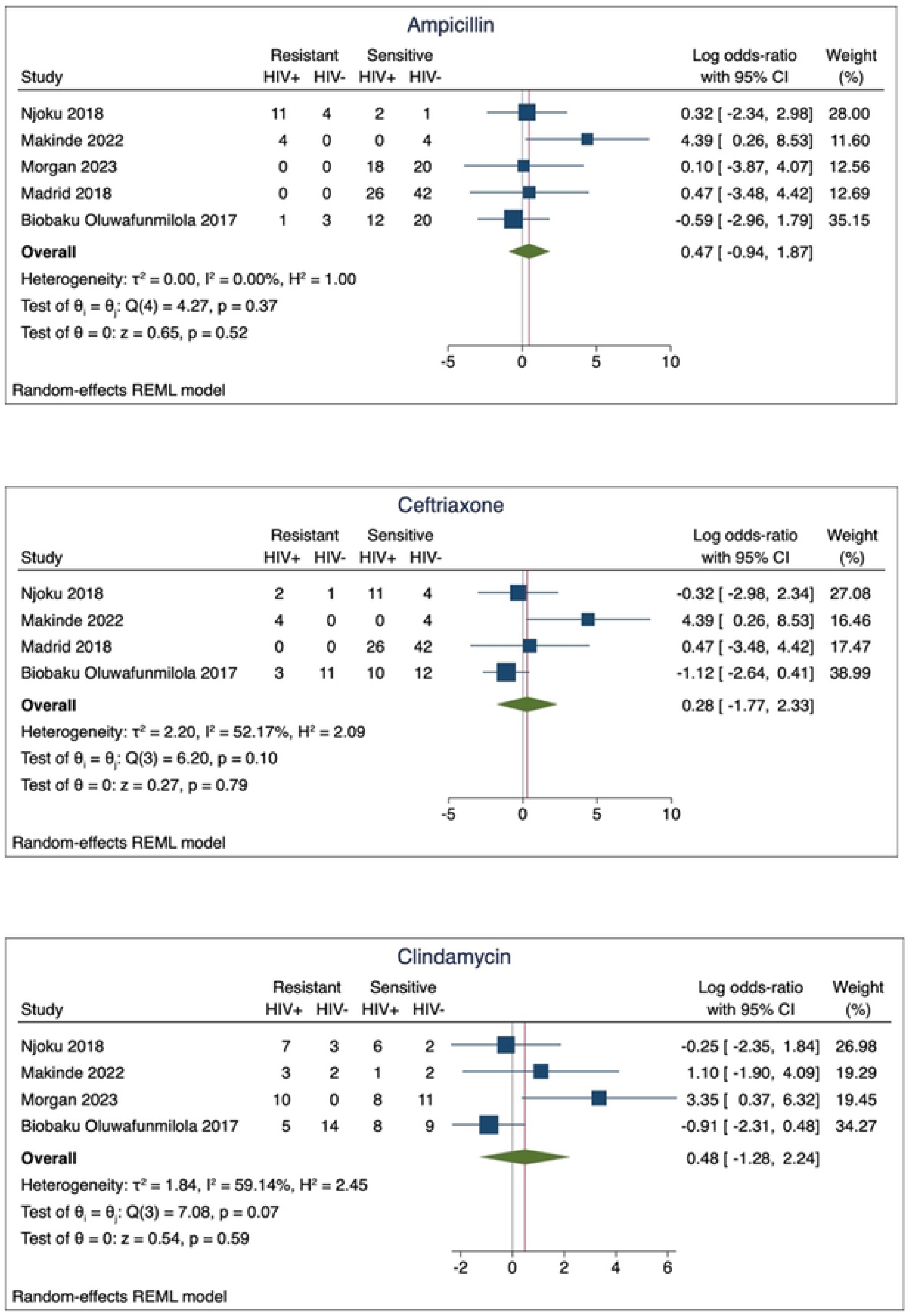
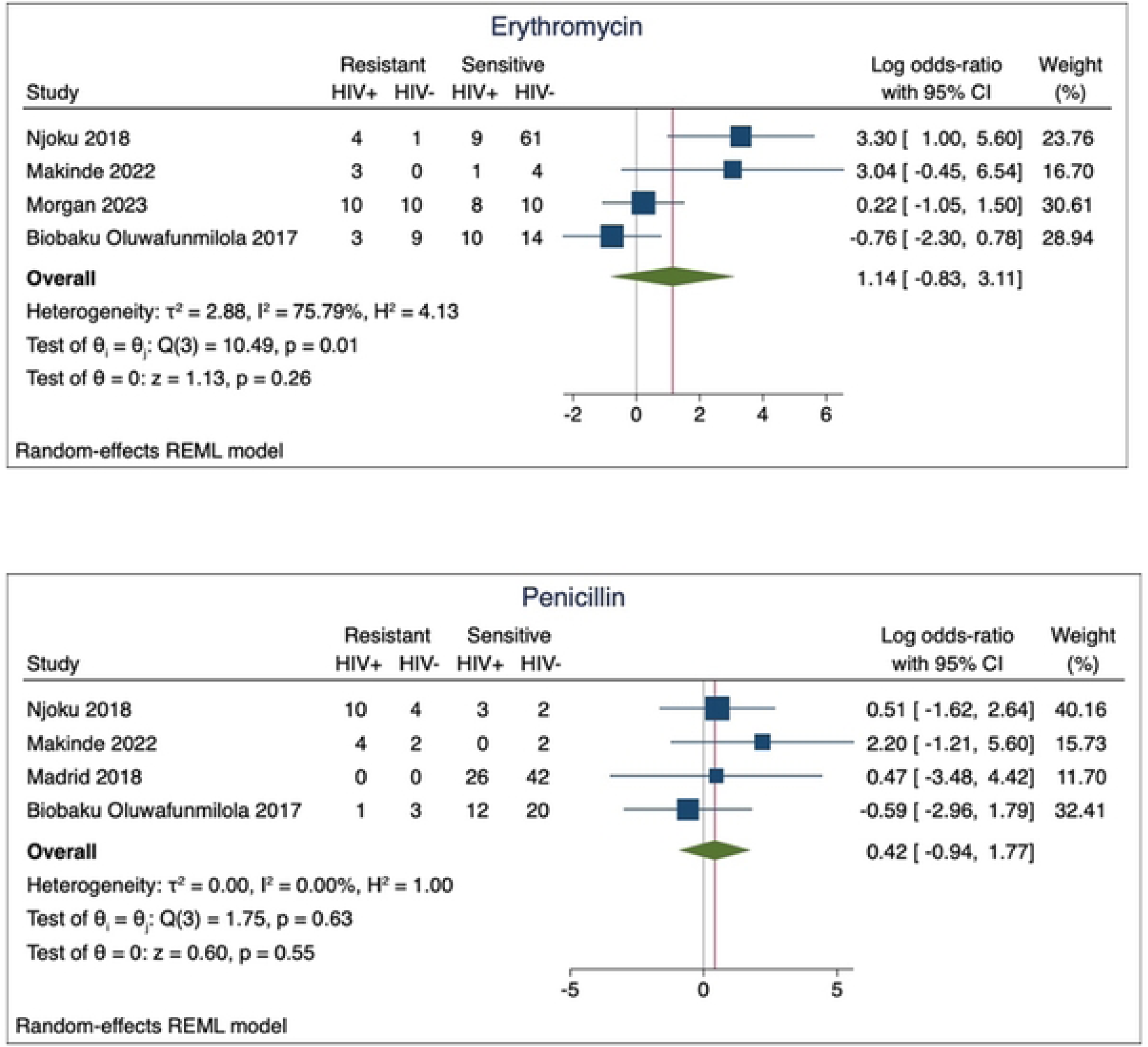
Pooled log odds of AMR among those with and without HIV, by antibiotics.

## Discussion

This systematic review and meta-analysis found no significant interactive effects between HIV and vaginal bacterial colonization on low birthweight in the included studies (OR=1.21, p=0.88). We found the odds of preterm birth among those with HIV and bacterial colonization was 2.64 times greater than those without HIV or colonization, albeit borderline non-significant (OR=2.64, *p*=0.05). The majority of studies tended toward increased odds of preterm birth with HIV and colonization, preterm birth remains a rare outcome, and even with pooling study results, we may have lacked the statistical power to adequately assess the effect. Unfortunately, due to both data availability limitations and limitations in the available data, we could not model the pooled effect on gestational age or birth weight outcomes as continuous variables, which would have improved statistical power. Future studies should report incidences of both continuous and categorical outcome measures to allow for a more nuanced assessment of this relationship.

We found no previous systematic review or meta-analysis which has assessed the potential interactive relationship between HIV status, vaginal bacterial colonization and LBW, though when assessed separately, systematic reviews and meta-analyses show that HIV-exposed neonates are at higher risk for GBS neonatal disease[44], and that low birth weight is more prevalent in neonates born from women living with HIV infection[45]. Our findings regarding preterm birth are similar to those from previous systematic review and meta-analyses that demonstrated increased odds of preterm birth among individuals with HIV and vaginal bacterial colonization compared to those without[45]. Results from Ravindran *et al*[34] study of a decreased risk of preterm birth in HIV-positive individuals with colonization differed from other studies. However, this finding may be attributable to the small sample size of HIV-positive participants in this study (n = 10) and the corresponding higher likelihood of type II error, as well as the fact that they did not recruit based on HIV status, and only retained incident cases of HIV in their originally HIV-negative cohort. Although there was only moderate heterogeneity among our included studies, varying patterns of pathogenic bacteria colonizing the lower genital tract, and the fact that *bacterial vaginosis* is a clinical syndrome caused by a group of bacterial species, rather than a single pathogen type, may have contributed to the observed heterogeneity in studies. Additionally, some methodological differences in the studies could contribute to our results, including differential methods and criteria for testing for bacterial colonization among groups with and without HIV, differing methods of defining preterm birth, and selection and inclusion criteria.

We also found that the overall odds of bacterial colonization did not differ significantly between those living with HIV and those without. This finding is congruent with a systematic review and meta-analysis conducted by Cools *et al.* which showed no difference between HIV status and rectovaginal colonization[44]. Our meta-analysis included 13 studies, however, the high degree of heterogeneity between studies limits the reliability of this estimate. Further, we found moderate to high risk of bias in most studies included in our analysis, due to methodological limitations. However, few of these studies were designed to assess the association between HIV status and bacterial colonization as a primary outcome. Future prospective cohort studies with strong selection and recruitment strategies, along with adequate confounding control in the analysis are necessary to confirm whether HIV status impacts the odds of bacterial colonization in pregnancy. Similarly, we did not observe a significant difference in the odds of antimicrobial resistance (AMR) among those living with HIV and those without. However, the sample sizes for each antibiotic were small, and due to a lack of individual-level data, we were unable to pool the analysis across antibiotic types, which limited statistical power. Although the specific antibiotics to which resistance exists is important to understand, future studies should also consider the inclusion of an analysis of whether pregnant individuals with colonization are resistant to one or more antibiotics, to allow an examination of the association between HIV status and resistance. In a meta-analysis conducted in the general population among people living with HIV, those with HIV were more likely to be colonized with resistant bacterial strains than people without and there was an increased risk of AMR in those living with HIV, across a range of bacterial pathogens and in multiple drug classes[46]. However, it remains unclear if similar patterns persist during pregnancy. Moreover, bacteria colonizing the lower genital tract in high-income countries (HICs) tend to be predominantly Gram-positive. In Low and Middle-Income Countries (LMICs), Gram-negative bacteria are predominant and are more highly associated with neonatal sepsis and mortality, highlighting a need for further research taking into account this geographical context.

Although this systematic review and meta-analysis presents up-to-date global evidence on the prevalence of vaginal bacterial colonization and pregnancy outcomes, it is not without limitations. The majority of the included studies are from Africa where the prevalence of HIV infection is high (60% of the HIV-infected population is in Africa). Comparatively, there is a lack of studies originating from Asia, Europe, and North and South America. The generalizability of the findings is therefore limited to African populations, as the overall sample is not representative of a global population. Further, most included studies recruited participants and/or laboratory samples through hospitals or from mothers who presented for routine ANC or delivery at tertiary hospitals. This could potentially skew the estimated prevalence towards urban settings and underestimate the associations that might exist in rural areas or smaller local health centres serving individuals of a more vulnerable socioeconomic status. The risk of antimicrobial-resistant organisms likely differs in these tertiary hospitals compared to rural settings and may be higher due to greater exposure to antimicrobials in people seeking care at tertiary hospitals.

One of the strengths of this study is the inclusion of studies examining direct vaginal colonization, whereas several previous studies evaluated rectal colonization only, meaning previous evidence may include pathogens that are not present in the lower genital tract, and thus may not influence pregnancy to the same extent[47, 48]. Although the large overall sample size of included studies in our review is a strength, the limited number of studies with complete data for inclusion in the meta-analyses of each outcome coupled with the moderate to high heterogeneity of studies limits the reliability of estimates, and our findings should be interpreted with caution. A further limitation of our findings is that there is likely some publication bias toward those with significant findings in the direction of elevated odds of adverse outcomes, which further limits the reliability of our findings.

The result of this meta-analysis indicates that there is still no clear evidence of whether HIV infection and bacterial colonization interact to affect low birth weight or preterm birth. There is still no global consensus on routine screening of pregnant women for lower genital tract pathogenic bacterial colonization, and the variations in methods used across the included studies are likely diminishing the reliability of our findings. Working toward a standardized vaginal bacterial screening during pregnancy, regardless of HIV status, could act to improve the identification of bacterial infections and potentially reduce any associated adverse outcomes. Further research conducted with large sample sizes, strong selection criteria, reliable and valid measurement, and adequate control for confounding variables, along with the assessment of birthweight and gestational age at delivery as continuous outcomes are desirable to strengthen the evidence in this area of research.

## Abbreviations

AMR: Antimicrobial Resistance
AMSTAR: Assessing the Methodological Quality of Systematic Reviews
ANC: Antenatal Care
CINAHL: Cumulative Index to Nursing and Allied Health Literature
CMA: Comprehensive Meta-Analysis
CI: Confidence Interval
HICs: High-Income Countries
HIV: Immunodeficiency Virus
IUGR: Intrauterine Growth restrictions
LBW: Low Birth Weight
LMICs: Low-Middle Income Countries
MESH: Medical Subject Headings
OR: Odds ratio
NIH: National Institutes of Health
NOS: New Castle-Ottawa Scale
PCR: Polymerase Chain Reaction
PRISMA: Preferred Reporting Items for Systematic Reviews and Meta-Analyses
SGA: Small for Gestation
SD: Standard Deviation
SSA: Sub-Saharan Africa

## Contributions

DM and KHC developed the protocol and were involved in the design, selection of study, data extraction, quality assessment, statistical analysis, results interpretation and development of the initial and final drafts of the manuscript; JS, JD, and MM conceived the project, designed the study and did several reviews of the manuscripts; DM, QG and KHC were involved in screening, quality assessment, data extraction, statistical analysis and revising subsequent manuscript drafts; PS and BK were involved in data analysis, synthesis and review of the manuscript. All authors read and approved the final draft of the manuscript.

## Ethics approval

Ethical approval was not required since this study relied solely on published data. However, permission for publication was sought from the CUHAS/BMC Research & Ethical Committee (CREC).

## Data availability

The data analyzed during the current systematic review and meta-analysis are available and made public (S3 Appendix).

## Acknowledgements

We would like to thank the staff of the University of Calgary, particularly Sophie Petit from the Indigenous, Local & Global Health (ILGH) office and Dianne Lorenzetti from the University Library, for their assistance with developing the search strategy. We also extend our gratitude to all the authors of the studies included in this systematic review and meta-analysis.

## References

1. Tumuhamye, J., Steinsland, H., Tumwine, J.K., Namugga, O., Mukunya, D., Bwanga, F., et al., Vaginal colonisation of women in labour with potentially pathogenic bacteria: a cross sectional study at three primary health care facilities in Central Uganda. BMC infectious diseases, 2020. 20(1): p. 98.

2. Sweeney, E.L., Kallapur, S.G., Gisslen, T., Lambers, D.S., Chougnet, C.A., Stephenson, S.A., et al., Placental infection with Ureaplasma species is associated with histologic chorioamnionitis and adverse outcomes in moderately preterm and late-preterm infants. The Journal of infectious diseases, 2016. 213(8): p. 1340–1347.

3. Juliana, N.C., Peters, R.P., Al-Nasiry, S., Budding, A.E., Morré, S.A. and Ambrosino, E., Composition of the vaginal microbiota during pregnancy in women living in sub-Saharan Africa: a PRISMA-compliant review. BMC pregnancy and childbirth, 2021. 21(1): p. 1–15.

4. Abdool Karim, S.S., Baxter, C., Passmore, J.A.S., McKinnon, L.R. and Williams, B.L., The genital tract and rectal microbiomes: their role in HIV susceptibility and prevention in women. Journal of the International AIDS Society, 2019. 22(5): p. e25300.

5. Charlier, C., O. Disson, and M. Lecuit, Maternal-neonatal listeriosis. Virulence, 2020. 11(1): p. 391–397.

6. Tumuhamye, J., Steinsland, H., Bwanga, F., Tumwine, J.K., Ndeezi, G., Mukunya, D., et al., Vaginal colonization with antimicrobial-resistant bacteria among women in labor in central Uganda: prevalence and associated factors. Antimicrobial Resistance & Infection Control, 2021. 10(1): p. 1–11.

7. Wang, P., Tong, J.J., Ma, X.H., Song, F.L., Fan, L., Guo, C.M., et al., Serotypes, antibiotic susceptibilities, and multi-locus sequence type profiles of Streptococcus agalactiae isolates circulating in Beijing, China. PLoS One, 2015. 10(3): p. e0120035.

8. Ozim, C.O., Mahendran, R., Amalan, M. and Puthussery, S., Prevalence of human immunodeficiency virus (HIV) among pregnant women in Nigeria: a systematic review and meta-analysis. 2023. 13(3): p. e050164.

9. Manyahi, J., Jullu, B.S., Abuya, M.I., Juma, J., Ndayongeje, J., Kilama, B., et al., *Prevalence of HIV and syphilis infections among pregnant women attending antenatal clinics in Tanzania*, *2011*. BMC Public Health, 2015. 15(1): p. 501.

10. De Goffau, M.C., Lager, S., Sovio, U., Gaccioli, F., Cook, E., Peacock, S.J., et al., Human placenta has no microbiome but can contain potential pathogens. Nature, 2019. 572(7769): p. 329–334.

11. Waldorf, K.M.A. and R.M. McAdams, Influence of infection during pregnancy on fetal development. Reproduction, 2013. 146(5): p. R151–R162.

12. Okwaraji, Y.B., Bradley, E., Ohuma, E.O., Yargawa, J., Suarez-Idueta, L., Requejo, J., et al., National routine data for low birthweight and preterm births: Systematic data quality assessment for United Nations member states (2000–2020). BJOG: An International Journal of Obstetrics & Gynaecology, 2023.

13. Ohuma, E.O., Moller, A.B., Bradley, E., Chakwera, S., Hussain-Alkhateeb, L., Lewin, A., et al., National, regional, and global estimates of preterm birth in 2020, with trends from 2010: a systematic analysis. The Lancet, 2023. 402(10409): p. 1261–1271.

14. Birhane Fiseha, S., Mulatu Jara, G., Azerefegn Woldetsadik, E., Belayneh Bekele, F. and Mohammed Ali, M., Colonization Rate of Potential Neonatal Disease-Causing Bacteria, Associated Factors, and Antimicrobial Susceptibility Profile Among Pregnant Women Attending Government Hospitals in Hawassa, Ethiopia. Infection and Drug Resistance, 2021. 14: p. 3159.

15. Kamgobe, E., Grote, S., Mushi, M.F., Wilson, D., Gandye, L., Bader, O., et al., Multi-drug resistant facultative pathogenic bacteria colonizing the vagina of pregnant women with premature rupture of membrane, Tanzania. East Africa Science, 2020. 2(1): p. 29–35.

16. Mitima, K.T., Ntamako, S., Birindwa, A.M., Mukanire, N., Kivukuto, J.M., Tsongo, K., Prevalence of colonization by Streptococcus agalactiae among pregnant women in Bukavu, Democratic Republic of the Congo. The Journal of Infection in Developing Countries, 2014. 8(09): p. 1195–1200.

17. Ernest, A.I., Ndaboine, E., Massinde, A., Kihunrwa, A. and Mshana, S., Maternal vaginorectal colonization by Group B Streptococcus and Listeria monocytogenes and its risk factors among pregnant women attending tertiary hospital in Mwanza, Tanzania. Tanzania Journal of Health Research, 2015. 17(2).

18. Shah, M., Aziz, N., Leva, N. and Cohan, D., Group B Streptococcus colonization by HIV status in pregnant women: prevalence and risk factors. Journal of women’s health, 2011. 20(11): p. 1737–1741.

19. Dauby, N., Adler, C., Miendje Deyi, V.Y., Sacheli, R., Busson, L., Chamekh, M., et al. *Prevalence, risk factors, and serotype distribution of group B Streptococcus colonization in HIV-infected pregnant women living in Belgium: a prospective cohort study*. in Open Forum Infectious Diseases. 2018. Oxford University Press US.

20. Page, M.J., McKenzie, J.E., Bossuyt, P.M., Boutron, I., Hoffmann, T.C., Mulrow, C.D., et al., The PRISMA 2020 statement: an updated guideline for reporting systematic reviews. International journal of surgery, 2021. 88: p. 105906.

21. Schiavo, J.H., PROSPERO: an international register of systematic review protocols. Medical reference services quarterly, 2019. 38(2): p. 171–180.

22. Wells GA, S.B., O’Connell D, Peterson J, Welch V, Losos M, et al., The Newcastle-Ottawa Scale (NOS) for assessing the quality of nonrandomised studies in meta-analyses. 2014.

23. Thompson, D.M. and Y.D. Zhao, Choosing statistical models to assess biological interaction as a departure from additivity of effects. arXiv preprint arXiv:2301.03349, 2023.

24. Madrid, L., Maculuve, S.A., Vilajeliu, A., Sáez, E., Massora, S., Cossa, A., et al., Maternal carriage of group B Streptococcus and Escherichia coli in a district hospital in Mozambique. The Pediatric infectious disease journal, 2018. 37(11): p. 1145–1153.

25. Nyemba, D.C., Peters, R.P., Medina-Marino, A., Klausner, J.D., Ngwepe, P., Myer, L., et al., Impact of aetiological screening of sexually transmitted infections during pregnancy on pregnancy outcomes in South Africa. BMC Pregnancy and Childbirth, 2022. 22(1): p. 194.

26. Gudza-Mugabe, M., Havyarimana, E., Jaumdally, S., Garson, K.L., Lennard, K., Tarupiwa, A., et al., Human immunodeficiency virus infection is associated with preterm delivery independent of vaginal microbiota in pregnant African women. The Journal of infectious diseases, 2020. 221(7): p. 1194–1203.

27. Njoku, C., C. Emechebe, and A. Agbakwuru, prevalence and determinants of anogenital colonization by Group B Streptococcus infection among HIV positive and negative women in Calabar, Nigeria. Int J Women’s Health Reprod Sci, 2017. 6(1): p. 11–7.

28. Price, J.T., Vwalika, B., France, M., Ravel, J., Ma, B., Mwape, H., et al., HIV-associated vaginal microbiome and inflammation predict spontaneous preterm birth in Zambia. Scientific Reports, 2022. 12(1): p. 8573.

29. Biobaku Oluwafunmilola, R., Olaleye Atinuke, O., Adefusi Olorunwa, F., Adeyemi Babalola, A., Onipede Anthony, O., et al., Group B streptococcus colonization and HIV in pregnancy: A cohort study in Nigeria. Journal of Neonatal-Perinatal Medicine, 2017. 10(1): p. 91–97.

30. Smullin, C.P., Green, H., Peters, R., Nyemba, D., Qayiya, Y., Myer, L., et al., Prevalence and incidence of Mycoplasma genitalium in a cohort of HIV-infected and HIV-uninfected pregnant women in Cape Town, South Africa. Sexually transmitted infections, 2020. 96(7): p. 501–508.

31. Short, C.E.S., Brown, R.G., Quinlan, R., Lee, Y.S., Smith, A., Marchesi, J.R., et al., Lactobacillus-depleted vaginal microbiota in pregnant women living with HIV-1 infection are associated with increased local inflammation and preterm birth. Frontiers in Cellular and Infection Microbiology, 2021. 10: p. 596917.

32. Makinde, O., B. Okusanya., and G. Osanyin., Group B Streptococcus vaginal colonization in pregnant women living with HIV infection: prevalence and antibiotic susceptibility at HIV referral centers in Lagos, Nigeria. The Journal of Maternal-Fetal & Neonatal Medicine, 2022. 35(25): p. 9098–9104.

33. Morgan, J.A., Hankins, M.E., Callais, N.A., Albritton, C.W., Vanchiere, J.A., et al., Group B Streptococcus rectovaginal colonization and resistance patterns in HIV-positive compared to HIV-negative pregnant patients. American Journal of Perinatology, 2023. 40(14): p. 1573–1578.

34. Ravindran, J., Richardson, B.A., Kinuthia, J., Unger, J.A., Drake, A.L., Osborn, L., et al., Chlamydia, gonorrhea, and incident HIV infection during pregnancy predict preterm birth despite treatment. The Journal of infectious diseases, 2021. 224(12): p. 2085–2093.

35. Gray, K.J., Kafulafula, G., Matemba, M., Kamdolozi, M., Membe, G. and French, N., *Group B Streptococcus and HIV infection in pregnant women, Malawi*, *2008*–2010. Emerging infectious diseases, 2011. 17(10): p. 1932.

36. El Beitune, P., Duarte, G., Maffei, C.M.L., Quintana, S.M., Rosa, A.C.J.D.S., Silva, E., et al., Group B Streptococcus carriers among HIV-1 infected pregnant women: prevalence and risk factors. European Journal of Obstetrics & Gynecology and Reproductive Biology, 2006. 128(1-2): p. 54–58.

37. Benedetto, C., Tibaldi, C., Marozio, L., Marini, S., Masuelli, G., Pelissetto, S., et al., Cervicovaginal infections during pregnancy: epidemiological and microbiological aspects. The Journal of Maternal-Fetal & Neonatal Medicine, 2004. 16(2): p. 9–12.

38. Foessleitner, P., Petricevic, L., Boerger, I., Steiner, I., Kiss, H., Rieger, A., et al., HIV infection as a risk factor for vaginal dysbiosis, bacterial vaginosis, and candidosis in pregnancy: A matched case-control study. Birth, 2021. 48(1): p. 139–146.

39. Govender, S., Theron, G.B., Odendaal, H.J. and Chalkley, L.J., Prevalence of genital mycoplasmas, ureaplasmas and chlamydia in pregnancy. Journal of Obstetrics and Gynaecology, 2009. 29(8): p. 698–701.

40. Zenebe, A., B. Eshetu, and S. Gebremedhin, Association between maternal HIV infection and birthweight in a tertiary hospital in southern Ethiopia: retrospective cohort study. Italian Journal of Pediatrics, 2020. 46: p. 1–9.

41. Joachim, A., Matee, M.I., Massawe, F.A. and Lyamuya, E.F., Maternal and neonatal colonisation of group B streptococcus at Muhimbili National Hospital in Dar es Salaam, Tanzania: prevalence, risk factors and antimicrobial resistance. BMC public Health, 2009. 9: p. 1–7.

42. Seale, A.C., Koech, A.C., Sheppard, A.E., Barsosio, H.C., Langat, J., Anyango, E., et al., Maternal colonization with Streptococcus agalactiae and associated stillbirth and neonatal disease in coastal Kenya. Nature microbiology, 2016. 1(7): p. 1–10.

43. Temmerman, M., Plummer, F.A., Mirza, N.B., Ndinya-Achola, J.O., Wamola, I.A., Nagelkerke, N., et al., Infection with HIV as a risk factor for adverse obstetrical outcome. Aids, 1990. 4(11): p. 1087–1094.

44. Cools, P., van de Wijgert, J.H., Jespers, V., Crucitti, T., Sanders, E.J., Verstraelen, H., et al., Role of HIV exposure and infection in relation to neonatal GBS disease and rectovaginal GBS carriage: a systematic review and meta-analysis. Scientific Reports, 2017. 7(1): p. 13820.

45. Xiao, P.L., Zhou, Y.B., Chen, Y., Yang, M.X., Song, X.X., Shi, Y., et al., Association between maternal HIV infection and low birth weight and prematurity: a meta-analysis of cohort studies. BMC pregnancy and childbirth, 2015. 15: p. 1–11.

46. Olaru, I.D., Tacconelli, E., Yeung, S., Ferrand, R.A., Stabler, R.A., Hopkins, H., et al., The association between antimicrobial resistance and HIV infection: a systematic review and meta-analysis. Clinical Microbiology and Infection, 2021. 27(6): p. 846–853.

47. Marando, R., Seni, J., Mirambo, M.M., Falgenhauer, L., Moremi, N., Mushi, et al., Predictors of the extended-spectrum-beta lactamases producing Enterobacteriaceae neonatal sepsis at a tertiary hospital, Tanzania. International Journal of Medical Microbiology, 2018. 308(7): p. 803–811.

48. Kovacs, D., Silago, V., Msanga, D.R., Mshana, S.E., Seni, J., Oravcova, K, et al., The hospital environment versus carriage: Transmission pathways for third-generation cephalosporin-resistant bacteria in blood in neonates in a low-resource country healthcare setting. Scientific Reports, 2022. 12(1): p. 8347.

